# The impact of the initial and 2^nd^ national COVID-19 lockdown on mental health in young people with and without pre-existing depressive symptoms

**DOI:** 10.1101/2021.11.03.21265861

**Authors:** Andrea Joensen, Stine Danielsen, Per Kragh Andersen, Jonathan Groot, Katrine Strandberg-Larsen

## Abstract

**Background:** The evidence on mental well-being and loneliness among young people during the initial lockdown is mixed, and little is known about the long-lasting impact of the sequential lockdowns. We examine changes in young people’s mental health from before to during the initial and second more prolonged lockdown, and whether women and those with pre-existing depressive symptoms were disproportionally impacted.

**Methods:** Participants reported on mental health indicators in an ongoing 18-year data collection in the Danish National Birth Cohort and in a COVID-19 survey, including 8 data points: 7 in the initial lockdown, and 1 year post. Changes in quality of life (QoL), mental well-being, and loneliness were estimated with random effect linear regressions on longitudinal data (N=32,985), and linear regressions on repeated cross-sections (N=28,579).

**Findings:** Interim deterioration in mental well-being and loneliness was observed during the initial lockdown, and only in those without pre-existing depressive symptoms. During the second lockdown, a modest deterioration was again observed for mental well-being and loneliness. QoL likewise only declined among those without pre-existing symptoms, where women showed a greater decline than men. QoL did not normalise during the initial lockdown and remained at lower levels during the second lockdown. These findings were not replicated in the repeated cross-sections.

**Interpretation:** Except for an interim decrease in mental health during lockdown, and only in those without pre-existing depressive symptoms, this study’s findings do not suggest a substantial detrimental impact of the lockdowns. Potential methodological differences in-between studies are a possible explanation for the mixed evidence.

**Funding:** The Velux Foundation

**Research in context:** *Evidence before this study:* We searched PubMed, PsycINFO, MedrXiv, and PsyArXiv with the terms (“Mental*” OR “Psychological*” OR “Emotional*”) AND (“Youth” OR “Young Adult*”) AND (“COVID*” OR “Coronavirus” OR “Lockdown*”) for articles published in English between January 1^st^ 2020 and October 1^st^ 2021. Included studies varied in terms of quality of data used but overall studies reported a detrimental impact of the lockdowns on young people’s mental health. However, the evidence on mental well-being and loneliness has shown to be inconsistent and with signs of resilience. Young people, women, and those with a pre-existing mental disorder have been identified as vulnerable subgroups, but only a few studies investigating mental health in individuals with a pre-existing mental disorder included a pre-lockdown measurement. The included studies also demonstrated that there is a gap in the evidence in understanding how mental health changed week by week, as well as the long-term impact over the course of the lockdowns.

*Added value of this study:* With longitudinal data, this study shows an interim impact of the initial and second lockdown on mental health during the COVID-19 pandemic in young individuals without pre-existing symptoms in Denmark. Since commencement of the initial lockdown, the levels of mental health returned to before levels, but one year after the initial lockdown, the levels were still lower than before lockdown in young people without pre-existing depressive symptoms. Young individuals with pre-existing depressive symptoms did not experience more detrimental impact of the lockdown, but rather indication of resilience or even improvements in mental health were observed. A disproportional impact of the lockdown on women compared to men was only observed for QoL, as women without pre-exiting depressive symptoms experienced a greater decline in QoL than men without pre-existing depressive symptoms. However, findings based on the repeated cross-sectional data did not show similar interim impact – but instead no – or clinically irrelevant impact. Thus, taken together our findings do not suggest a substantial lasting impact of the lockdowns on mental health among young individuals.

*Implications of all available evidence:* A great majority of earlier studies suggest that the lockdowns due to the COVID-19 pandemic have had substantial detrimental impact on mental health, and that women and those with a pre-existing mental disorder constitute vulnerable subgroups. However, these studies vary considerably in terms of method applied. Our findings emphasise the importance of the use of different data setups, as well as methodology applied for the investigation of mental health. More studies based on high-quality data used in different settings are needed to fully understand the impact of the lockdowns on young people’s mental health, including potential disproportional impact on vulnerable subgroups.

## Introduction

The COVID-19 pandemic became a global reality in early 2020 with enormous impact on society and daily living. Like many other countries worldwide, lockdowns, quarantine requirements and recommendations, social restrictions, and physical distancing were implemented in Denmark in March 2020 to mitigate the spread of the virus. The Danish lockdown demanded all public employees with no critical function to work from home, closing of national borders, schools, day-care centers, sports facilities, and restaurants. Moreover, private companies were strongly recommended to let their employees work from home.^1^ This initial lockdown was eased during late spring, but then gradually reinforced during the autumn 2020 in response to rising numbers of cases and deaths attributed to COVID-19. Mid December, a 2^nd^ national strict lockdown was implemented that lasted to March 2021, from which a gradually reopening began. Several studies have documented acutely deteriorations of mental health and increased loneliness during the initial lockdowns compared with pre-pandemic periods.^2–5^ However, according to the Lancet’s COVID-19 Commission Mental Health Task Force the evidence up to December 2020 regarding the impact of COVID-19 on well-being and loneliness was inconsistent and with signs of resilience.^6^ Several studies have also investigated whether women and young people have been disproportionally impacted by the lockdown.^2,3,7–18^ The majority of these studies did identify women and young people as vulnerable subgroups.^2,3,7–9,14,15,17,18^ Further, patient organisations, case stories, and health professionals have raised concerns about marked worsening of pre-existing mental disorders during the lockdowns. Studies support that people with pre-existing mental disorders were impacted more detrimentally by the lockdown.^7,8^ Contrary, other studies, all with before and during measures of mental health, have shown that the changes in mental health were minimal or even slightly improved in people with severe and chronic mental disorders, whereas the deteriorations in mental health were among people without pre-existing mental disorders.^15,19,20^

Research on the mental health impact of COVID-19 and related lockdowns is evolving fast, but not many studies are based on high-quality data and only a few studies in young people include a before measure and up to several measures during the lockdown. Additionally, we lack knowledge on how the full picture will unfold, and how the sequential and prolonged lockdowns have impacted young people’s mental health. The aim of this study was to quantify changes in quality of life (QoL), mental well-being, and loneliness in young people following and through the initial lockdown, and during the second and more prolonged lockdown. We further examined whether women and individuals with pre-existing depressive symptoms were disproportionally impacted by the lockdowns, as we hypothesised these to be most vulnerable.

## Methods

### Participants

In the mid-nineties, the nationwide national birth cohort, the Danish National Birth Cohort (DNBC) was established, into which 30% of children born in Denmark in 1996-2003 were enrolled.^21^ Longitudinal data exist from prenatal life unto early adulthood collected in the latest data sweep, the 18-year data collection (DNBC-18). The DNBC-18 was initiated in 2016 and will be completed ultimo 2021. Further information is available: www.dnbc.dk. To document the public health impact of the national COVID-19 lockdown, we invited participants to complete a COVID-19 survey. Only participants who had earlier provided either their private mail or phone number were invited. Further eligibility criteria were an active security number and not having withdrawn participation. The initial COVID-19 survey, determined wave 1, was launched in the 3^rd^ week of the initial lockdown, Figure 1. All participants who responded within a week were re-invited to up to six subsequent consecutive online surveys, i.e. wave 2-7.^1^ Approximately one year later, i.e. April/May 2021, all participants with identical eligible criteria were re-invited to wave 8 of the COVID-19 survey.

**Figure 1.**
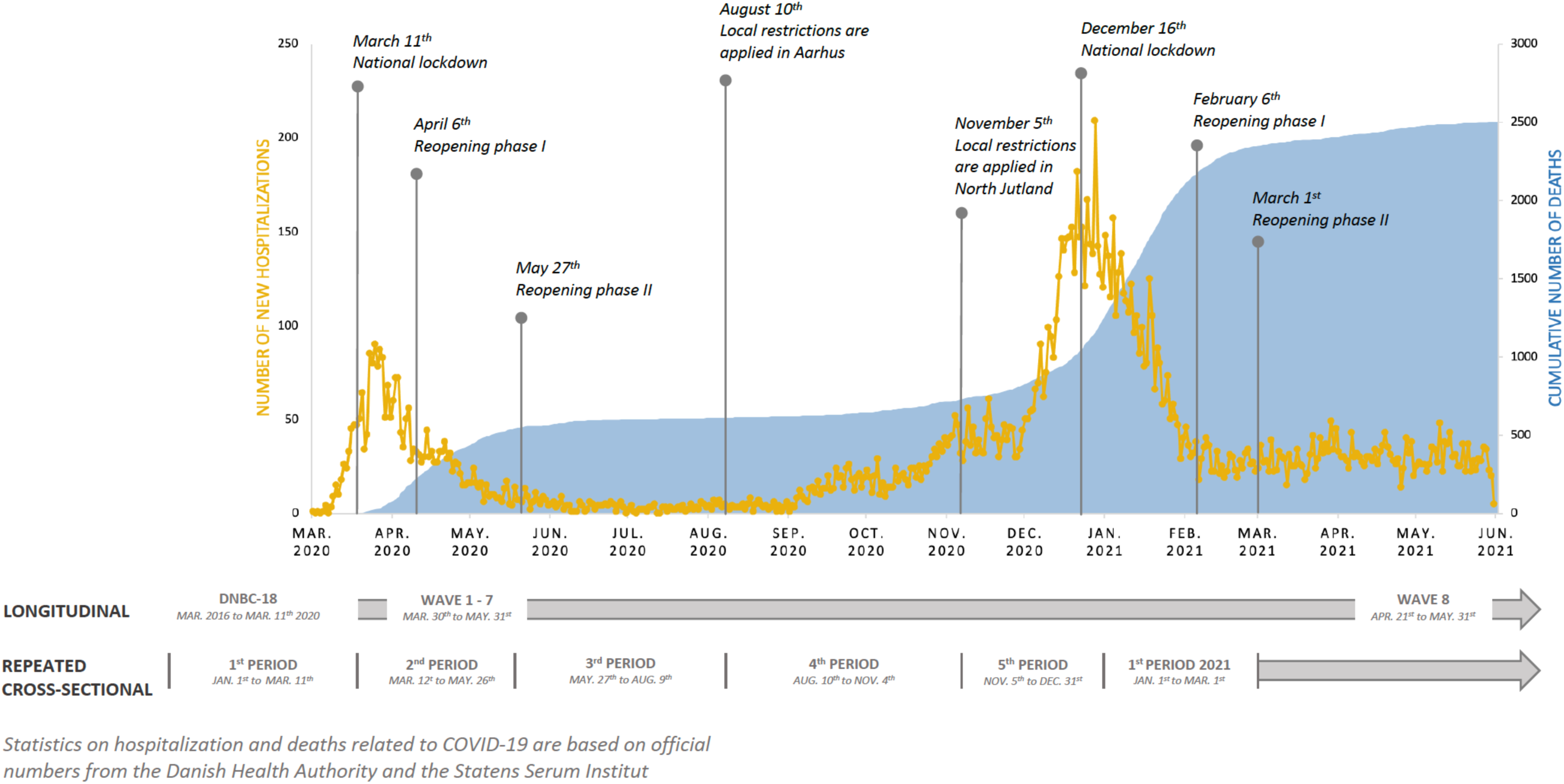
The data set up presented according to the development of the COVID-19 pandemic and following lockdowns in Denmark.

The populations, in the present study, were restricted to participants with information on household-socio-occupational status, maternal age at childbirth, parity, and maternal smoking collected during pregnancy. In the analyses including the DNBC-18 and the COVID-19 survey, we further restricted our population to those eligible for the DNBC-18 before the initial lockdown, Figure S1. In total, 32,985 participants had complete data in the DNBC-18. Of these, 7,431 and 8,808, respectively participated in wave 1 and wave 8 of the COVID-19 survey, Figure S1. We also estimated the year-to-year and seasonal variation in mental health by utilising the DNBC-18 collected in 2018 to March 2021 (N=28,579) as repeated cross-sections divided into year and five periods, Figure S2. The periods reflected the initiation, reopening, and reinforcements of the lockdowns in Denmark, Figure 1.

### Primary mental health outcome measures

Primary mental health measures in this study, include two widely used measures of QoL and mental well-being, that have shown good reliability and validity.^22,23^ The measure of loneliness in this study, was only based on a single item.

#### QoL

We used an adaptation of the Cantril Ladder scale, in which respondents rate their life from 0 for the worst life to 10 for the best possible life, to measure QoL.^22^ This adaption of the Cantril Ladder scale is widely used internationally among adolescents and has shown good reliability and convergent validity with other emotional well-being measures.^22^

#### Mental well-being

We used the 7-item Short Warwick-Edinburgh Mental Well-Being Scale (SWEMWBS),^24^ which is a validated instrument, also in a Danish age-appropriate sample, to measure mental well-being.^23^ The response-option for each item is a five-point likert scale. Thus, the total scale ranges from 7-35, with higher values indicating better well-being. In the DNBC-18 and wave 8, the items referred to the previous two weeks, whereas in wave 1-7 of the COVID-19 survey, the items were rephrased to the specific week. A 1-point change on the scale is considered to represent a clinically meaningful change.

#### Loneliness

In the DNBC-18, participants were asked ‘How often do you feel lonely?’ with the response options ‘Never’, ‘Occasionally’, ‘Often’, ‘Very often’ or ‘Do not know’ (excluded). In the COVID-19 survey the item on loneliness was: ‘In the last week, how often have you felt lonely?’ with response options: ‘Seldom or not at all (less than 1 day)’, ‘Some or a little (1-2 days)’, ‘Occasionally or often (3-4 days)’ or ‘Most of the time (5-7 days)’. The two highest, i.e. at least ‘Often’ and ‘Occasionally or often (3-4 days)’ were categorised as lonely, and otherwise participants were categorised as not lonely.

## Measure of pre-existing depressive symptoms

### Pre-existing depressive symptoms

Measure of pre-existing depressive symptoms was assessed in the DNBC-18 by the Major Depression Inventory (MDI). The MDI is a validated instrument referring to feelings in the past two weeks and ranging from 0-50, with higher scores indicating more severe depression.^25^ Pre-existing depressive symptoms was categorised as scoring ≥26, and we further categorised severity of depressive symptoms: severe (31-50), moderate (26-30), mild (21-25), and no depression (0-20).^25^

### Covariates

Participants reported their current educational enrolment and housing composition in the DNBC-18. We also included information on gender, age, household-socio-occupational status, maternal age at childbirth, parity, and maternal smoking collected during pregnancy. These covariates were categorised as shown in Table S1.

### Statistical analysis

To account for differential attrition, we estimated inverse probability weights (IPW) by logistic regressions with having data as outcome and the following predictors: gender, household-socio-occupational status, maternal age at childbirth, parity, and maternal smoking collected during pregnancy. Separate analyses were performed for each data point and on the appropriate baseline population, Figure S1 and S2. Age at time of wave 1 was additionally included in the models for the COVID-19 waves. These IPWs were included in all analyses including the specific data points.

Using the longitudinal data, we estimated the mean changes with corresponding 95% confidence intervals (CI), in QoL, mental well-being, and proportion of change in loneliness by subtracting the pre-lockdown measurement from the lockdown measurement. For the periods in 2018-2021 in the repeated cross-sectional setup, we estimated the mean of QoL and mental well-being and the proportion being lonely, with corresponding 95% CI. These calculations were stratified on gender and pre-existing depressive symptoms, respectively. Next, we performed random effects linear regressions on the longitudinal data and linear regressions on the repeated cross-sectional data to estimate the changes in mean QoL and mental well-being, as well as the proportion being lonely, respectively, from before to during lockdown. In the longitudinal setup, we examined the changes from before to during lockdown by including wave 1-8. We contrasted before with during lockdown in a model with a binary variable for lockdown, gender, and pre-existing depressive symptoms. To test for disproportional impact of the lockdown among women vs. men and young people with vs. without pre-existing depressive symptoms, we gradually expanded the models with interactions. First with the interaction between lockdown and gender and then the interaction with pre-existing symptoms. Interactions were included if disproportional impacts were observed for at least one of the mental health outcomes. Additionally, we investigated whether severity of pre-existing depressive symptoms mattered by including the four categories of severity in the analyses. To examine whether the impact of the lockdown varied across the waves of the COVID-19 survey, we exchanged the binary lockdown variable with a variable indicating wave 1-8. Lastly, we restricted the before measure in the longitudinal analyses to participants who completed DNBC-18 in year 2019 and period 2 or 3, to address whether our results were biased by seasonal variation or the time gap between the pre- and during lockdown measurement.

In the repeated cross-sectional setup, the during lockdown period was defined as the second period in 2020 and onwards. We started out testing for interaction between lockdown and period and omitted it if insignificant. We subsequently examined the disproportional impact of lockdown on gender and depressive symptoms by including interaction terms as described above. We performed these analyses unadjusted and adjusted for household-socio-occupational status, maternal age at childbirth, maternal smoking collected during pregnancy, educational enrolment, and household composition.

The analyses were performed with SAS Software, version 9.4 (SAS Institute, North Carolina, US) using the commands proc survey means, proc mixed/GLM and applying the weight statement for IPW and random statement for random effect.

## Results

Slightly more than half of the participants were 18-20 years of age during the initial lockdown. Moreover, more women than men participated in the DNBC-18, and were undertaking education, living with parents, and from educated households. In the COVID-19 survey, seven out of ten participants were women in wave 1 and wave 8, while this proportion slightly increased in wave 2-7, Table S1. Moreover, participants in wave 2-7 were more often without pre-existing depressive symptoms, under education, living with parents, from educated households, nulliparous, non-smoking, and older mothers than participants within the DNBC-18 and wave 1 and 8, Table S1.

Before lockdown, women and young people with depressive symptoms reported lower mental health than men and those without depressive symptoms, Table S2. Deteriorations in QoL, mental well-being, and loneliness was observed in the strictest phase of the initial lockdown. Mental well-being and loneliness reached the before levels during the initial lockdown, while the QoL never normalised, Figure 2. One year post the initial lockdown (wave 8), reflecting the easing up after the second more prolonged lockdown, the QoL and mental well-being were at same levels as observed early during the initial lockdown. Loneliness was only slightly increased at wave 8 compared to before. Similar patterns were seen for women and men, while it was young people without pre-existing depressive symptoms who experienced the deteriorations.

**Figure 2.**
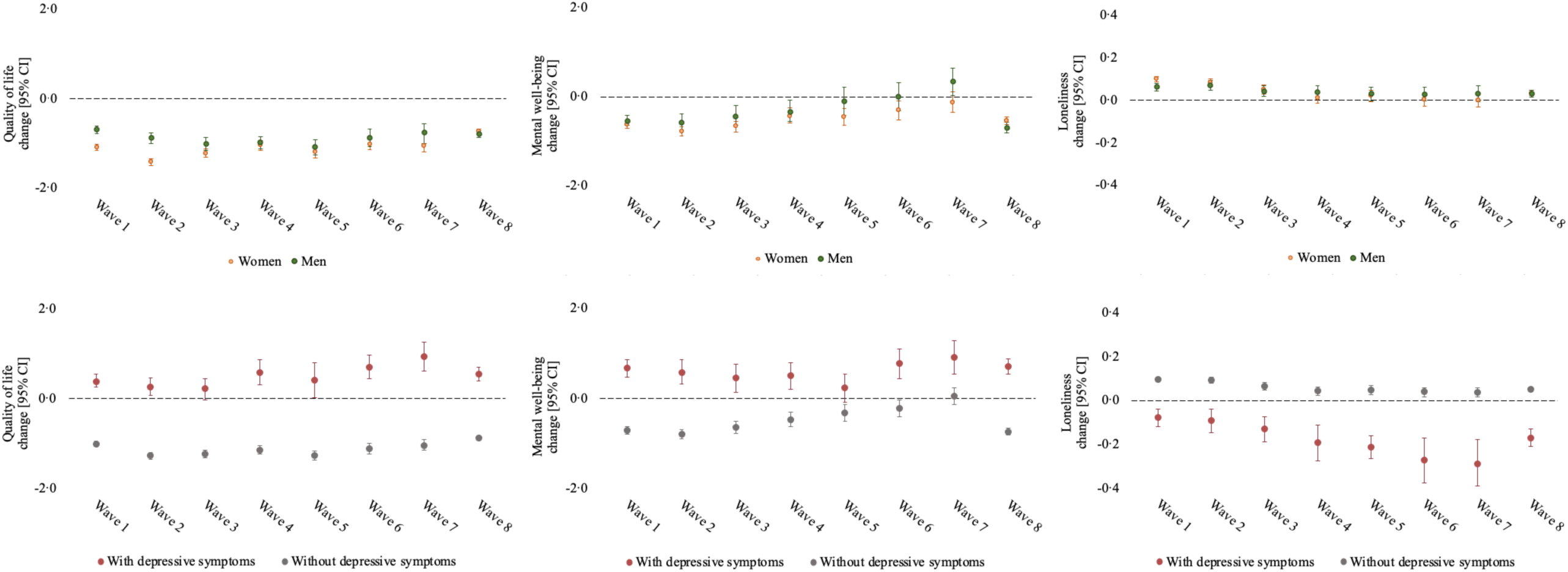
Mean change from pre- to during lockdown [95% CI] in QoL and mental well-being, and proportion of change in loneliness stratified by gender and pre-existing depressive symptoms, respectively (longitudinal setup)

For QoL, the lockdown had a disproportional impact on gender and pre-existing depressive symptoms groups, Figure 3. The biggest decline in QoL following lockdown was observed for women without pre-exiting depressive symptoms [-1·12, 95% CI:-1·17;-1·07], while QoL declined -0·85 point [95% CI:-0·90;-0·80] among men without pre-existing depressive symptoms. The lowest level of QoL was at wave 2-5 for both groups, and then slightly increased later in the initial lockdown. The level of QoL was still lower compared with before lockdown for both groups in spring 2021. Contrary, the QoL improved in young people with pre-existing depressive symptoms, especially in men. For mental well-being and loneliness, it was likewise people without-pre-existing depressive symptoms who experienced the deteriorations, while those with pre-existing depressive symptoms improved. Women and men without pre-existing depressive symptoms were not disproportionally impacted by the lockdown, as the drop in mental well-being was -0·63 point [95% CI:-0·71;-0·55] for women and -0·59 point [95% CI:-0·67;-0·50] for men and the proportion feeling lonely increased by 8·0% [95% CI:7·0;9·0%] for women and 6·0% [95% CI:5·0;7·0%] for men. The deteriorations in well-being and loneliness were greatest early in the initial lockdown. In spring 2021, the overall changes in mental well-being and loneliness from before lockdown were approximately the same as the change observed during the initial lockdown. When investigating the degree of pre-existing symptoms, the deteriorations were greatest for the no depressive symptom group and greatest improvements were seen for the severe group, Figure S3. Restricting the before measure to data collected in spring and summer 2019 did not change the overall conclusion, Figure S4.

**Figure 3.**
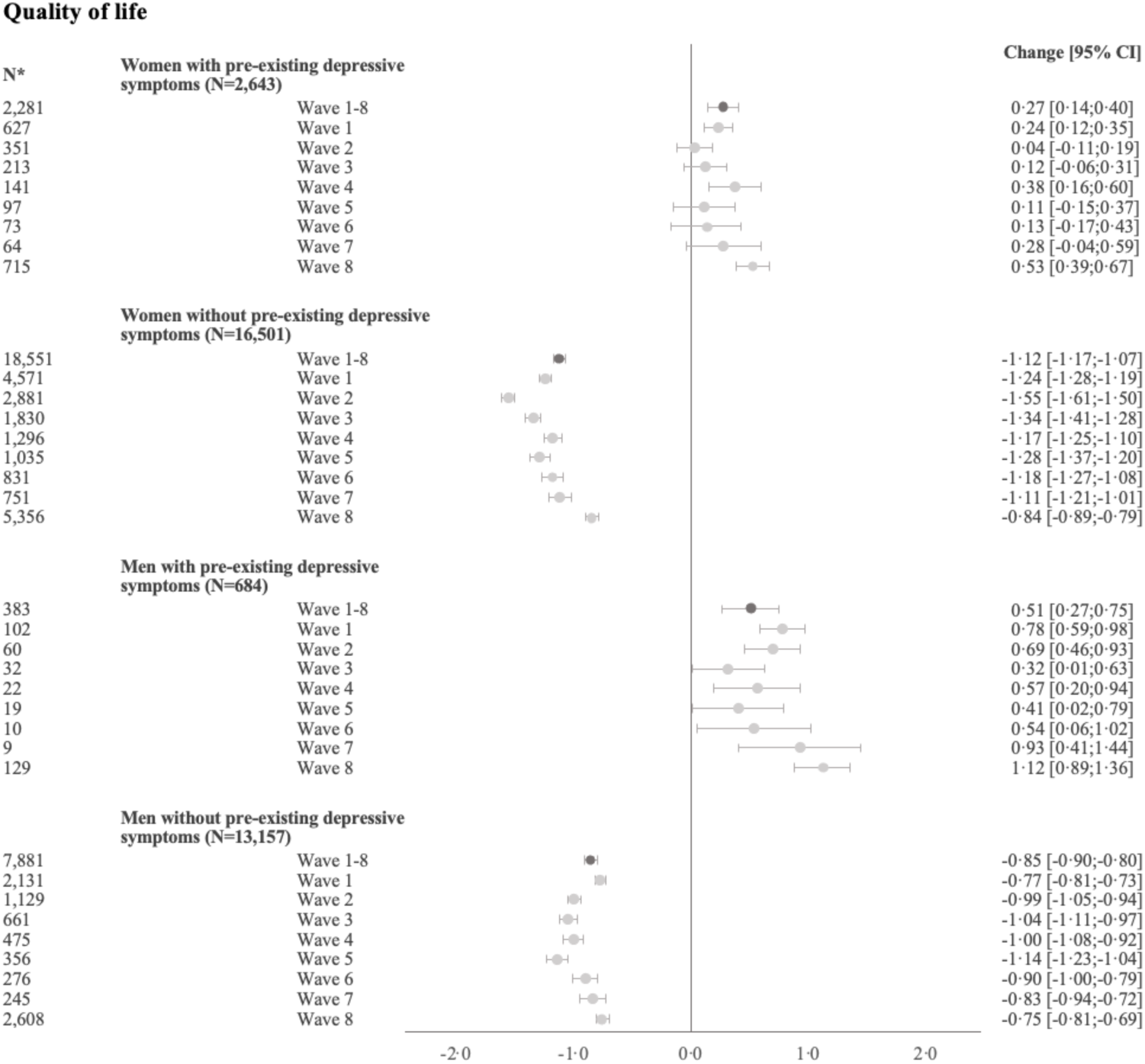
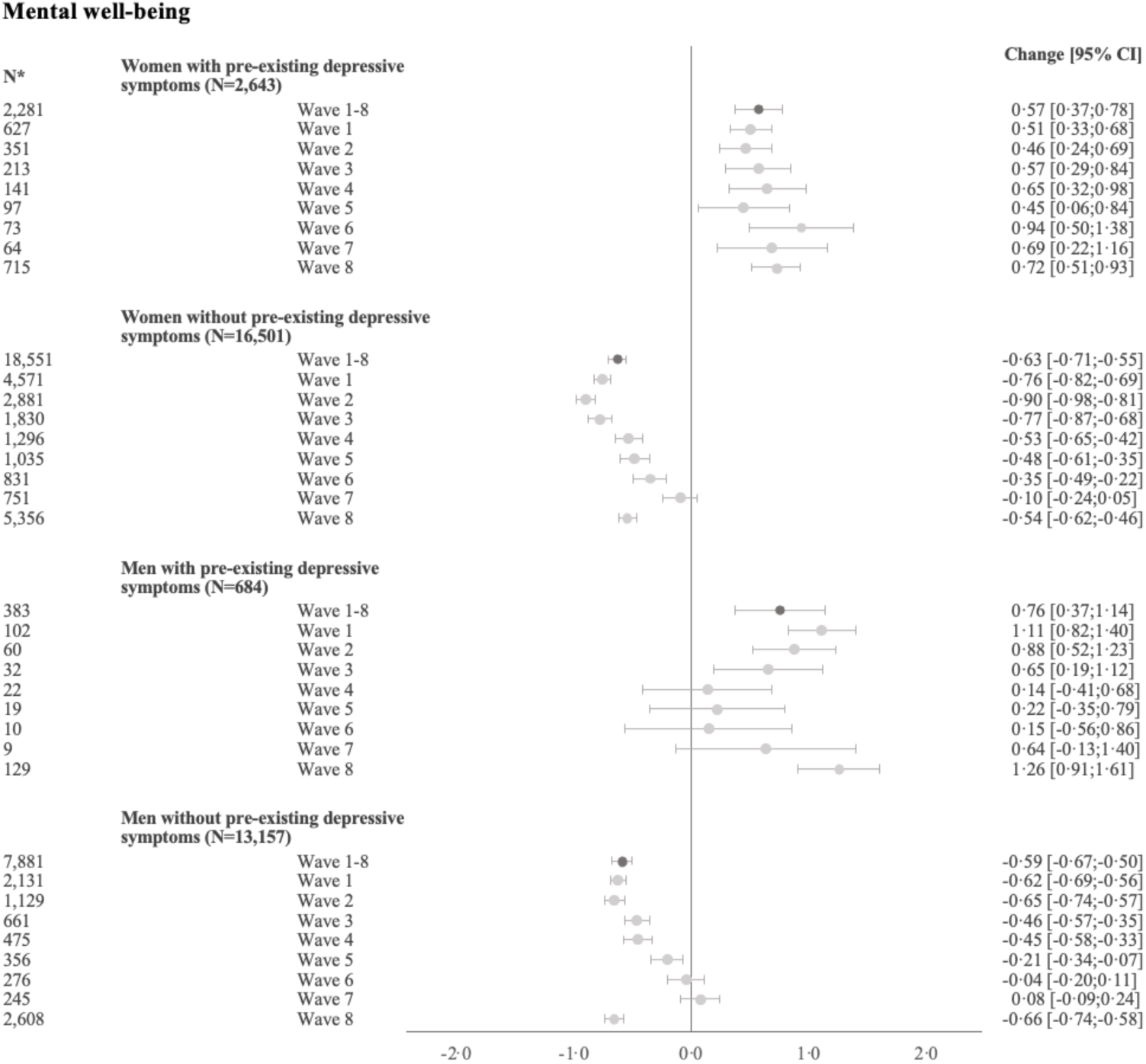
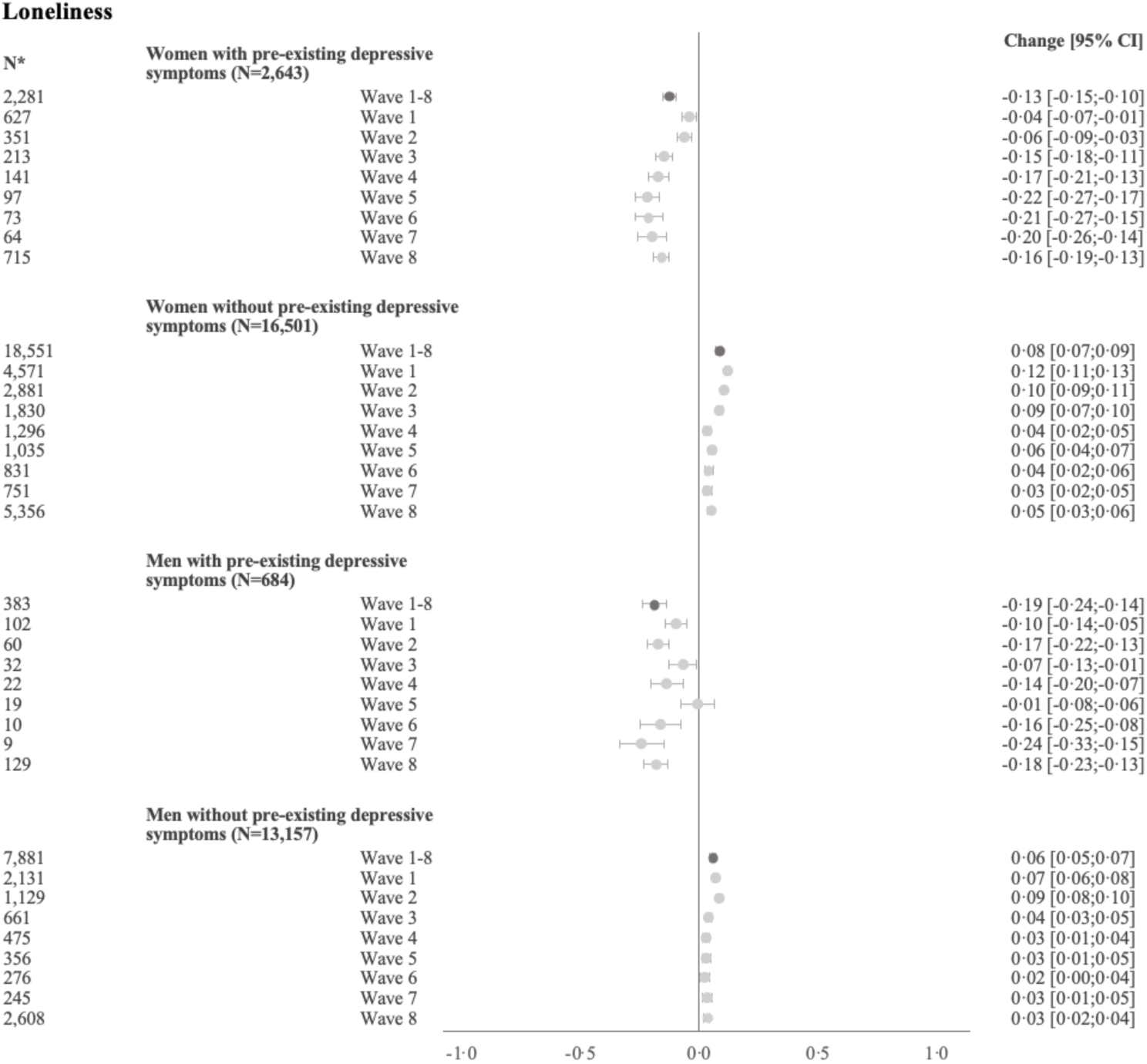
Regression of changes in QoL, mental well-being, and loneliness from pre- to during lockdown (longitudinal setup) *Repeated measures Random effect estimates and 95% CI presented (N=32,985) (Total number including repeated measures N=62,081) All models were weighted by IPW baseline population 1, Figure S2 (N=67,346) p-value for interaction between lockdown, gender, and pre-existing depressive symptoms (wave 1-8): QoL (p<0·001), mental well-being (p<0·001), and loneliness (p<0·001)

No clear changes were observed in QoL, mental well-being, or loneliness when analysing the repeated cross-sections, Figure 4. In the regression analyses, the period effect was equal before and after lockdown. For QoL and loneliness, the lockdown had a minor disproportional impact on the depressive symptoms groups, as the QoL only dropped slightly [-0·17, 95% CI:-0·22;-0·13] in people without depressive symptoms, Figure 5. Similarly, a minor improvement was observed in mental well-being among people without depressive symptoms [0·09, 95% CI:0·01;0·17], whereas no improvements were seen in those with depressive symptoms. Contrary, a 3·0% [95% CI:1·0;6·0%] increase in loneliness was observed in young people with depressive symptoms, while no changes were observed among people without depressive symptoms.

**Figure 4.**
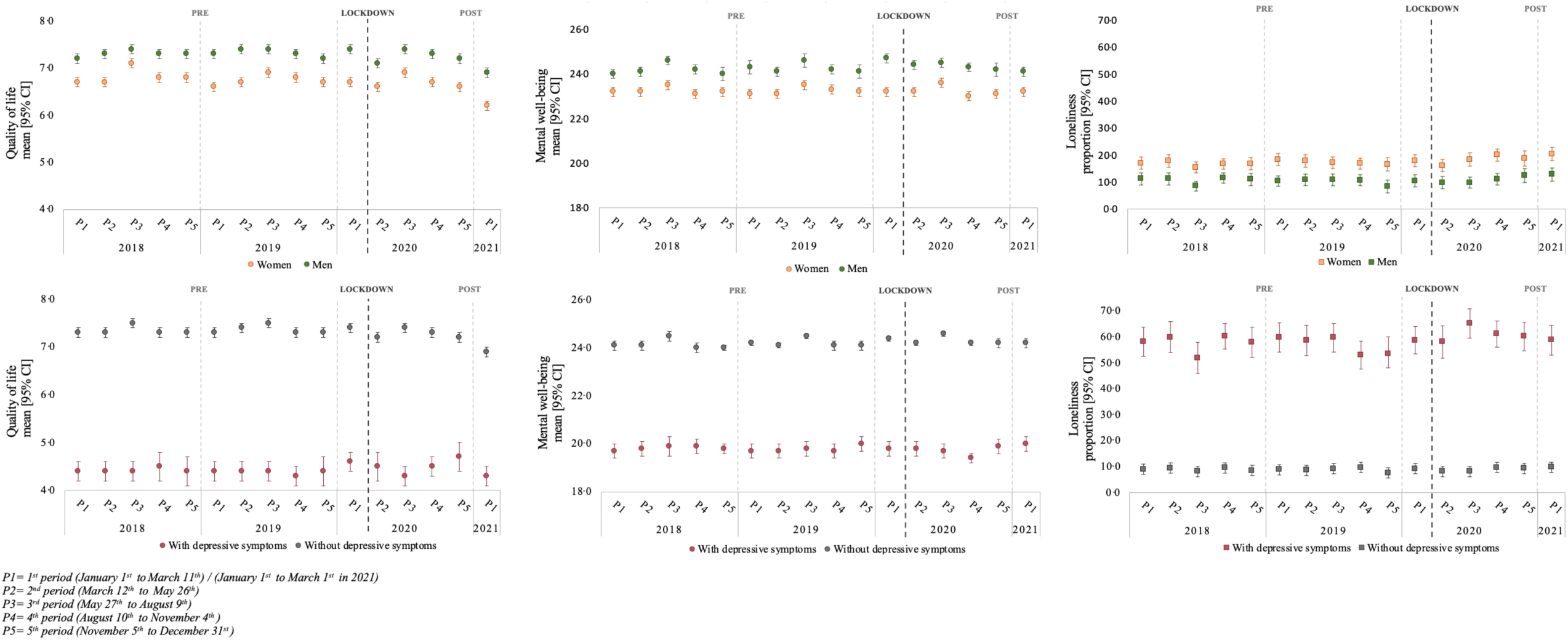
Mean/ proportion [95% CI] of QoL, mental well-being, and loneliness stratified by gender and depressive symptoms, respectively (repeated cross-sectional setup)

**Figure 5.**
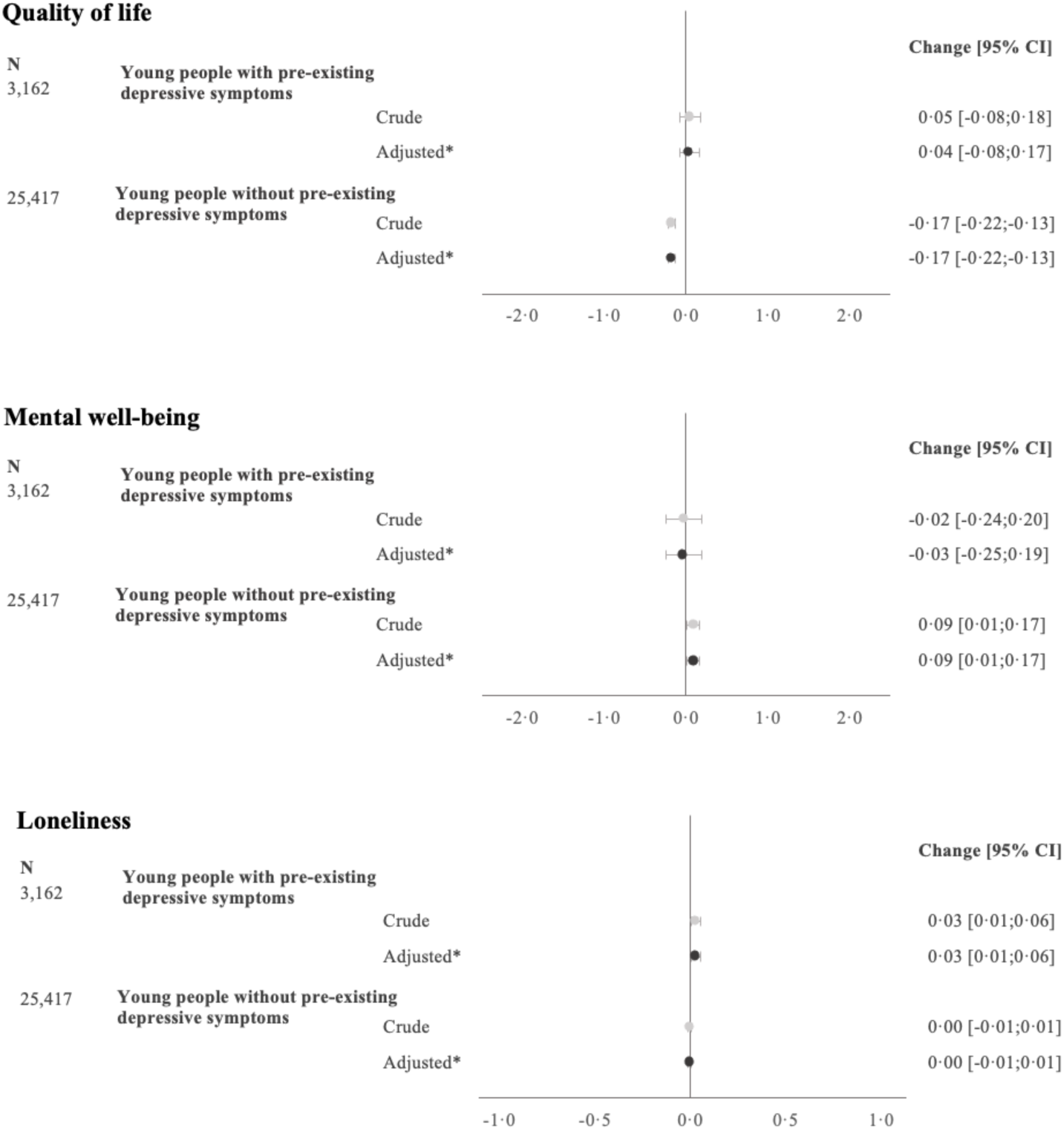
Regression of changes in QoL, mental well-being, and loneliness from pre- to during lockdown (repeated cross-sectional setup) Crude and adjusted estimates and 95% CI are presented (N=28,579) *Adjusted for gender, period, household socio-occupational status, maternal age at childbirth, maternal smoking during pregnancy, educational enrolment, and housing composition. All models were weighted by IPW baseline population 2, Figure S2 (N=58,638) p-value for interaction between lockdown and depressive symptoms: QoL (p<0·001), mental well-being (p=0·3365), and loneliness (p<0·001)

## Discussion

Within the longitudinal setup, this study demonstrates an interim deterioration in mental well-being and loneliness during the initial lockdown, and only in young people without pre-existing depressive symptoms. During the gradual re-opening of the second lockdown, the mental well-being was equivalent to early in the initial lockdown, while the proportion of loneliness was at levels during the reopening of the initial lockdown, thereby only slightly increased. QoL likewise only declined following lockdown among young people without pre-existing symptoms, but women had a bigger decline in QoL than men. QoL did not normalise among young people without pre-existing symptoms during the initial lockdown and remained at lower levels in spring 2021. Overall, these deterioration in mental health seemed rather modest, and were not replicated in repeated cross-sectional setup. Thus, taken together our findings do not suggest a substantial lasting impact of the lockdowns on mental health among young individuals.

The longitudinal data allow us to quantify the week-to-week variation in impact across the entire span of the initial lockdown. In contrast, in the repeated cross-sections, the initial lockdown is represented by one longer period. Mental well-being and loneliness seemed to normalise during the gradual reopening of the initial lockdown, and this might explain why we only observe deteriorations in QoL in young people without depressive symptoms in the cross-sectional analyses. Feelings of loneliness and social isolation since the implementation of the initial lockdown have been very dynamic and responsive to the government’s actions.^1,18^ Additionally, in the COVID-19 survey it was explicitly stated that the aim was to investigate how the COVID-19 pandemic impacted our living, which was not the case in the ongoing DNBC-18, where COVID-19 was not mentioned and no adaptions were made. The DNBC participants are regularly invited to complete age specific follow ups. Thus, it is likely, however untestable, that the participants in the DNBC-18 deliberately compensated, so their responses reflected their overall health and not solely their current lockdown situation.

Studies that have suggested a decline in young people’s mental health are cross-sectional or do not include a pre-lockdown measurement.^9,11–14,16^ Most of the earlier findings do also indicate that women were more detrimentally impacted by the lockdown than men,^2,3,7–9,14,15,17,18^ but for individuals with a pre-existing mental disorder, the findings are mixed.^5,7,8,10,11,15,18–20^ One possible explanation for the mixed findings can be the methodological differences. Regression to the mean is of concern, and longitudinal studies in which adjustment for pre-lockdown mental health was performed, documented greater deteriorations following the lockdown in individuals with pre-existing mental health disorders.^7,8^ Contrary, studies without adjustment for baseline values, as in our study, document slightly improved or unchanged mental health following lockdown among those with a pre-existing mental disorder.^10,15,19,20^ Observational studies examining change and adjusting for baseline values can lead to bias in the direction of the cross-sectional association between pre-existing depressive symptoms and the pre-lockdown measure of mental health.^26–28^ Individuals with pre-existing depressive symptoms scored substantially lower on QoL, well-being, and loneliness, and thus the association between pre-existing depressive symptoms and these mental health indicators reverses after adjustment for the pre-lockdown levels.^29^

Findings from the longitudinal setup showed that young people with pre-existing depressive symptoms experienced a resilience or an improvement during lockdown, for which there could be multiple possible explanations. The lockdown and the social isolation might have given individuals with depressive symptoms more calmness, as the new circumstances were in line with their normal daily life. Young people without pre-existing depressive symptoms however showed a deterioration in mental health which might represent a normal fear in response to an unpredicted crisis such as the COVID-19 pandemic. The results from wave 8 suggest a more long-term impact of the lockdown, as the levels of mental health were lower than before lockdown. However, wave 8 was during the gradual re-opening of the second lockdown, and if we presume it likely to follow same responsiveness as during the initial lockdown, we find it plausible it would have been higher if measured later in the re-opening phase. Although this study did not show any deteriorations among young people with pre-existing depressive symptoms, it is important to clarify that the mental health of these individuals was and remained systematically worse compared to those without pre-existing depressive symptoms.

### Strengths and limitations

Strength’s worth highlighting, is the tandem use of longitudinal data on individuals aged 18-24 and repeated samples of individuals aged 18 years originating from the same baseline population. Moreover, our data collection during the initial lockdown included up to 7 measurements spanning the reopening phases, as well as one measurement subsequent to a second and more prolonged lockdown. The repeated cross-sections allow us to quantify seasonal variations and is only vulnerable to attrition if the participation in DNBC-18 systematically changed over year of birth or season, as opposed to the longitudinal setup which is more vulnerable to attrition due to loss to follow-up. We attempted to reduce bias from differential attrition by inverse probability weighting. The validity of this method relies on a correctly specified model including all relevant predictors for loss to follow-up, which cannot be assumed.

Interpretation of our findings likewise deserves consideration of a few limitations. In the longitudinal setup, the baseline data was collected at age 18 years and three months for all participants, whereas the participants’ ages during lockdown was 18-24 years. Thus, the timespan between the before and during lockdown measurement was greater for the older participants. For older participants, changes in mental health may be underestimated, since the pre-lockdown measurement represent a younger age than the follow-up measures, and on average reporting on mental health instruments improves with age.^17^ As an attempt to preclude this, a sensitivity analyses was conducted to ensure that any observed deterioration in mental health was likely attributable to the lockdown. Finally, the DNBC has previously been shown to be healthier and more often from households with higher occupational levels than the background population, and our population was mainly living with their parents and studying.^30^ Thus, the findings from this study cannot necessarily be generalised to all 18–24-year-olds or to other countries.

In summary, the findings from the longitudinal setup did reveal a modest intermittent deterioration in mental health during the initial lockdown in young individuals without depressive symptoms prior to lockdown, as well as lower QoL and well-being 1 year post the initial lockdown than before lockdown. The mental health of young individuals with depressive symptoms prior to lockdown did not show similar deteriorations but remained unchanged or even slightly improved. Only for QoL, women without pre-existing depressive symptoms experienced a greater decline compared than men. These findings interpreted simultaneously with the findings from the repeated cross-sections, indicating no or clinical meaning less change do not support a substantial and lasting impact of the lockdown on the mental health in young individuals.

## Data Availability

According to European law (General Data Protection Regulation), data containing potentially identifying or sensitive personal information are restricted. However, for academic researcher, data could be available on request via DNBC

## Acknowledgment

The Danish National Birth Cohort (DNBC) was established with a significant grant from the Danish National Research Foundation. Additional support was obtained from the Danish Regional Committees, the Pharmacy Foundation, the Egmont Foundation, the March of Dimes Birth Defects Foundation, the Health Foundation, and other minor grants. The DNBC Biobank has been supported by the Novo Nordisk Foundation and the Lundbeck Foundation. Follow-up of mothers and children has been supported by the Danish Medical Research Council (SSVF 0646, 271-08-0839/06-066023, O602-01042B, 0602-02738B), the Lundbeck Foundation (195/04, R100-A9193), The Innovation Fund Denmark 0603-00294B (09-067124), the Nordea Foundation (02-2013-2014), Aarhus Ideas (AU R9-A959-13-S804), a University of Copenhagen Strategic Grant (IFSV 2012) and the Danish Council for Independent Research (DFF – 4183-00594 and DFF – 4183-00152). We thank Professor Lau Caspar Thygesen for a valuable discussion of the analytical model and interpretation of results.

