## Supplementary for "The impact of the initial and 2^nd^ national COVID-19 lockdown on mental health in young people with and without pre-existing depressive symptoms"

**Table S1.** Characteristics of baseline population and participants (unweighted)

| | Baseline [a] | 18-year [b] | 18-year [c] | Wave 1 | Wave 2 | Wave 3 | Wave 4 | Wave 5 | Wave 6 | Wave 7 | Wave 8 |
|---|---|---|---|---|---|---|---|---|---|---|---|
| | | N=32,985 | N=28,579 | N=7,431 | N=4,421 | N=2,736 | N=1,934 | N=1,507 | N=1,190 | N=1,069 | N=8,808 |
| | (%) | (%) | (%) | (%) | (%) | (%) | (%) | (%) | (%) | (%) | (%) |
| **Gender** | | | | | | | | | | | |
| Women | 49 | 58 | 59 | 70 | 73 | 75 | 74 | 75 | 76 | 76 | 69 |
| Men | 51 | 42 | 41 | 30 | 27 | 25 | 26 | 25 | 24 | 24 | 31 |
| **Pre-existing depressive symptoms [d]** | | | | | | | | | | | |
| With | .. | 10 | 11 | 10 | 9 | 9 | 8 | 8 | 7 | 7 | 10 |
| Without | .. | 90 | 89 | 90 | 91 | 91 | 92 | 92 | 93 | 93 | 90 |
| **Education [d,e]** | | | | | | | | | | | |
| Low/Medium/High education | .. | 80 | 79 | 86 | 88 | 88 | 88 | 89 | 89 | 90 | 86 |
| Other education | .. | 15 | 15 | 10 | 9 | 9 | 9 | 8 | 8 | 7 | 10 |
| Not student | .. | 5 | 6 | 4 | 3 | 3 | 3 | 3 | 3 | 3 | 4 |
| **Housing composition [d]** | | | | | | | | | | | |
| Living with both parents | .. | 62 | 62 | 65 | 67 | 69 | 70 | 71 | 70 | 70 | 67 |
| Living with one parent | .. | 30 | 30 | 28 | 27 | 25 | 25 | 24 | 25 | 25 | 26 |
| Living without parents | .. | 8 | 8 | 7 | 6 | 6 | 5 | 5 | 5 | 5 | 7 |
| **Age at invitation for wave 1** | | | | | | | | | | | |
| 18-20 years | 49 | 48 | .. | 55 | 54 | 53 | 52 | 52 | 52 | 53 | 55 |
| > 20 years | 51 | 52 | .. | 45 | 46 | 47 | 49 | 48 | 48 | 47 | 45 |
| **Household socio-occupational status** | | | | | | | | | | | |
| High educational level | 23 | 25 | 25 | 28 | 28 | 28 | 29 | 30 | 30 | 30 | 28 |
| Medium educational level | 31 | 32 | 33 | 35 | 36 | 37 | 37 | 37 | 36 | 37 | 34 |
| Skilled worker | 28 | 27 | 26 | 23 | 22 | 20 | 20 | 20 | 20 | 20 | 24 |
| Unskilled worker | 14 | 13 | 12 | 11 | 11 | 11 | 10 | 10 | 10 | 9 | 11 |
| Student | 2 | 2 | 2 | 3 | 3 | 3 | 3 | 2 | 3 | 3 | 3 |
| Outside of labor marked | 1 | 1 | 1 | 1 | 1 | 1 | 1 | 1 | 1 | 1 | 0 |
| Unclassifiable | <1 | <1 | <1 | <1 | <1 | <1 | <1 | <1 | <1 | <1 | 0 |
| **Maternal age at childbirth** | | | | | | | | | | | |
| < 25 years | 8 | 7 | 7 | 6 | 5 | 4 | 4 | 4 | 4 | 4 | 5 |
| 25-29 year | 35 | 35 | 36 | 35 | 35 | 36 | 34 | 35 | 36 | 36 | 36 |
| 30-34 year | 39 | 40 | 39 | 40 | 41 | 41 | 42 | 41 | 40 | 40 | 41 |
| ≥ 35 years | 17 | 18 | 18 | 19 | 19 | 19 | 19 | 19 | 20 | 20 | 18 |
| **Parity** | | | | | | | | | | | |
| 0 | 46 | 47 | 50 | 51 | 51 | 51 | 51 | 52 | 54 | 54 | 51 |
| 1 | 38 | 37 | 35 | 35 | 34 | 34 | 34 | 35 | 33 | 32 | 34 |
| ≥ 2 | 16 | 16 | 15 | 15 | 15 | 15 | 14 | 14 | 14 | 14 | 15 |
| **Smoking during pregnancy** | | | | | | | | | | | |
| Not smoking | 74 | 76 | 76 | 79 | 79 | 81 | 82 | 83 | 83 | 83 | 78 |
| Stopped smoking | 9 | 9 | 9 | 8 | 8 | 7 | 7 | 7 | 7 | 7 | 9 |
| 1-10 g daily | 13 | 12 | 11 | 10 | 10 | 9 | 8 | 8 | 7 | 8 | 10 |
| > 10 g daily | 4 | 4 | 3 | 3 | 3 | 3 | 3 | 3 | 3 | 2 | 3 |

[a] Baseline population 1 (n=67,346) and baseline population 2 (n=58,638). The characteristics did not vary in the two baseline populations. The presented characteristics are for baseline population 1.
[b] DNBC-18 used in the longitudinal analyses, Figure S1.
[c] Participants in DNBC-18 in 2018 to 2021, Figure S2.
[d] Data collected in DNBC-18.
[e] Low/Medium/High education is defined as high school to higher education (> 4 years) and other education is defined as primary school or other education on same level

**Table S2.** Mean QoL and mental well-being, as well as the proportion feeling lonely in DNBC-18 in the longitudinal setup (N=32,985), stratified by gender and depressive symptoms, respectively

| | Gender | | | | Depressive symptoms | | | |
|---|---|---|---|---|---|---|---|---|
| | Women N=19,144 | | Men N=13,841 | | With N=3,327 | | Without N=29,658 | |
| | Mean [95% CI] | | Mean [95% CI] | | Mean [95% CI] | | Mean [95% CI] | |
| QoL | 6·79 | [6·77;6·82] | 7·37 | [7·33;7·39] | 4·41 | [4·34;4·48] | 7·36 | [7·34;7·38] |
| Mental well-being | 23·24 | [23·19;23·28] | 24·27 | [24·21;24·31] | 19·78 | [19·69;19·86] | 24·18 | [24·14;24·21] |
| Loneliness | 16·89 | [16·36;17·43] | 10·31 | [9·80;10·82] | 57·35 | [55·69;59·00] | 8·94 | [8·61;9·27] |

**Figure S1.** Flowchart of the longitudinal data with the baseline populations and response rates in respectively the DNBC-18 and the COVID-19 survey wave 1-8

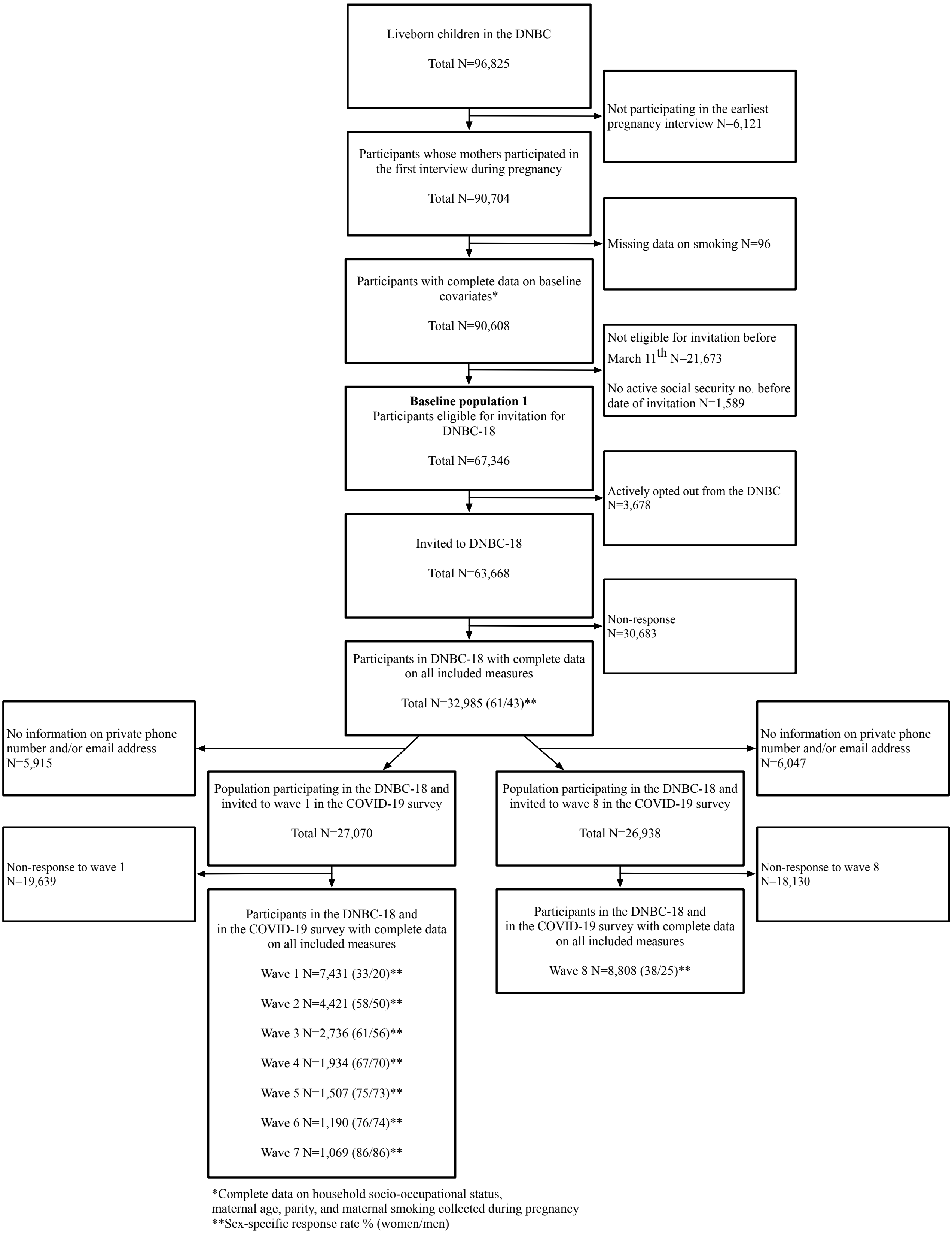


*Complete data on household socio-occupational status, maternal age, parity, and maternal smoking collected during pregnancy
**Sex-specific response rate % (women/men)

**Figure S2.** Flowchart of the baseline population for the DNBC-18 in year 2018-2021

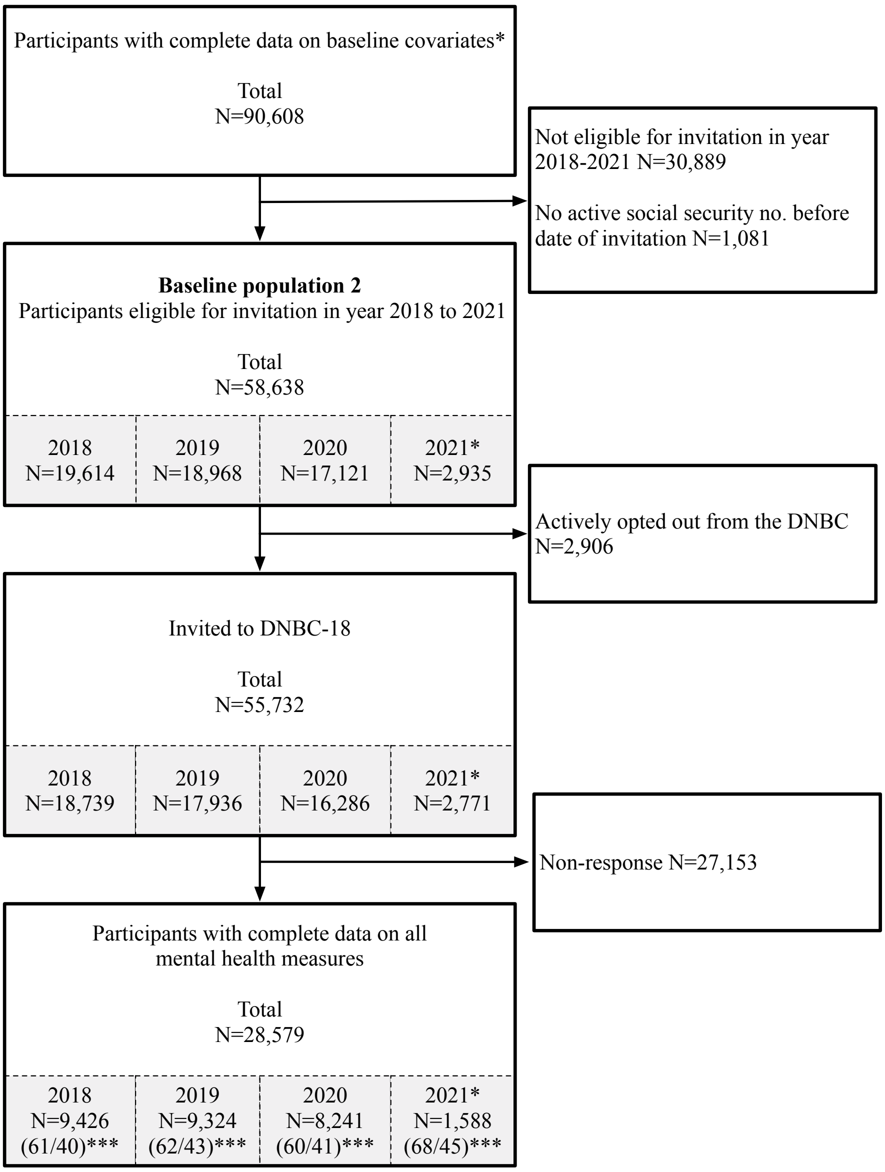


*Year 2021 only included data up until March 2021
**Complete data on household socio-occupational status, maternal age, parity, and maternal smoking collected during pregnancy
***Sex-specific response rate % (women/men)

**Figure S3**. Regression of changes in QoL, mental well-being, and loneliness from pre- to during lockdown. Pre-existing depressive symptoms divided into severe, moderate, mild, and no symptoms (longitudinal setup)

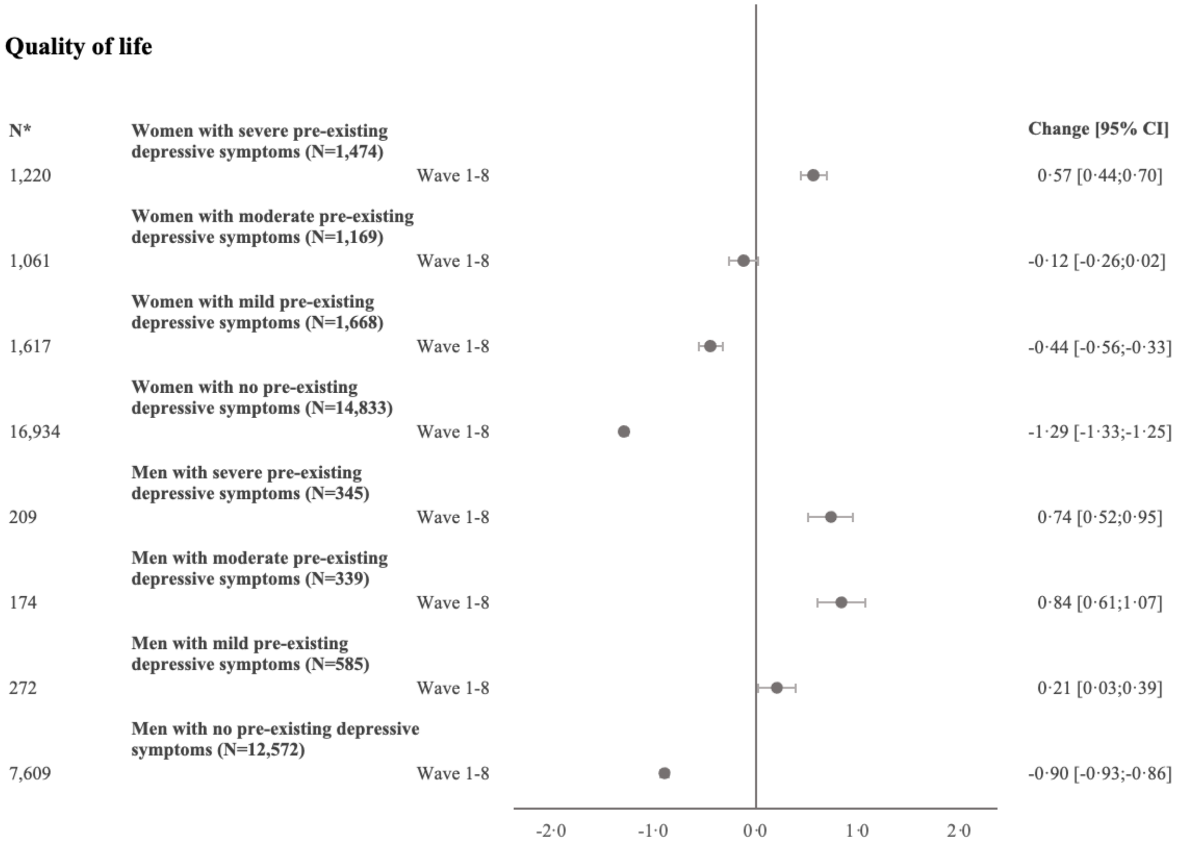


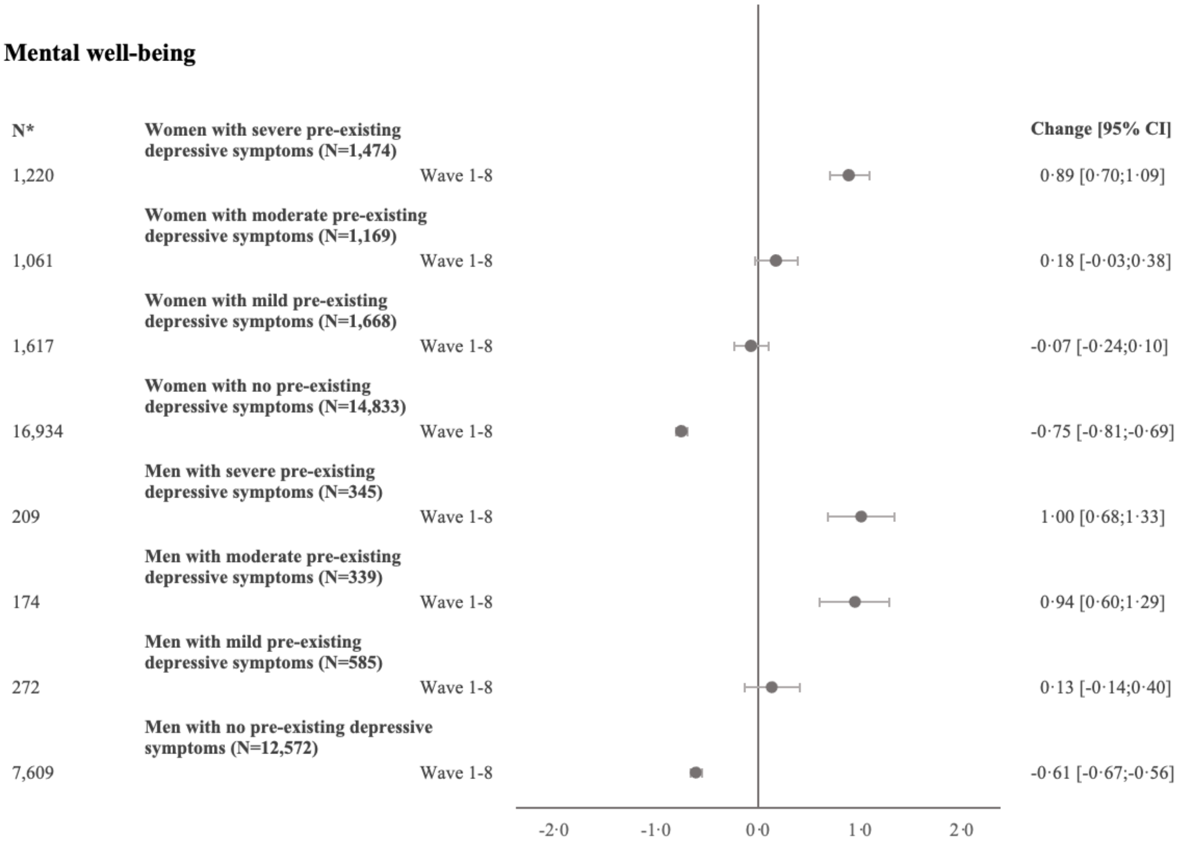

**Loneliness**

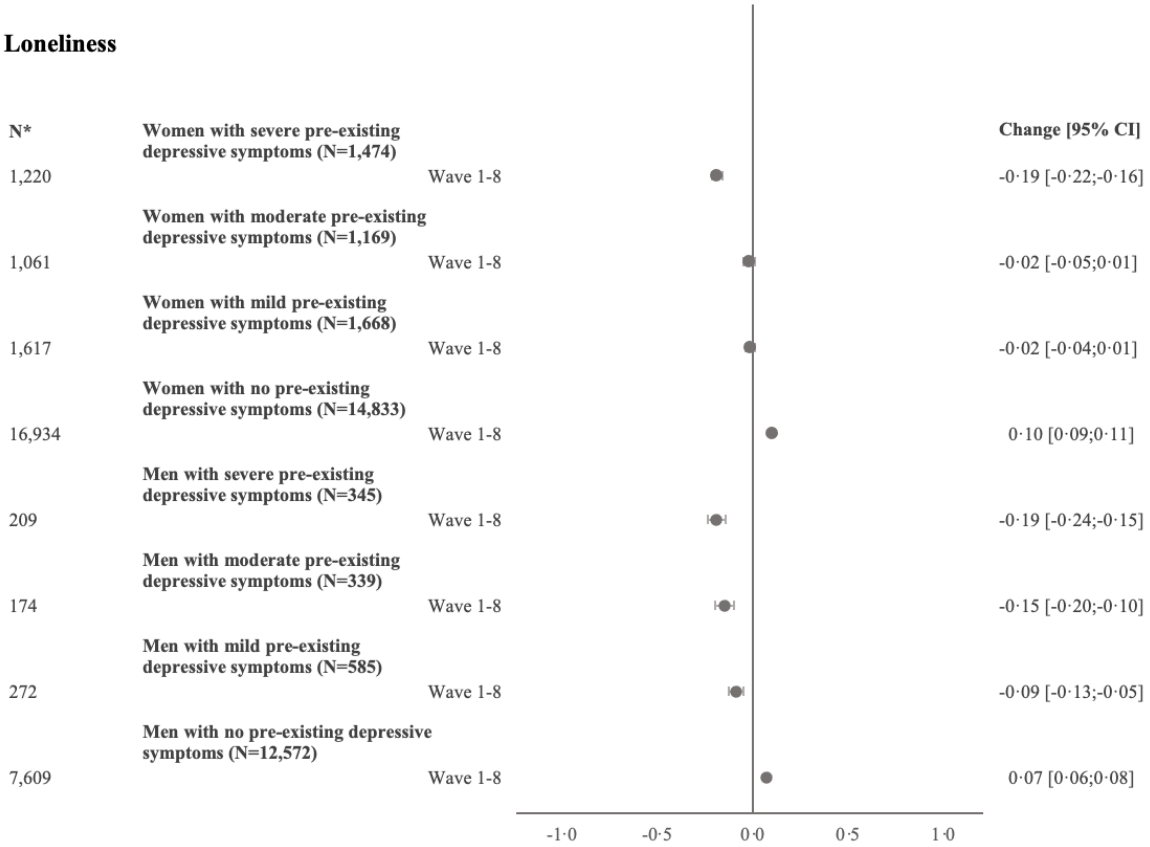


*Repeated measures
Random effect estimates and 95% CI presented (N=32,985)
(Total number including repeated measures N=62,081)
All models were weighted by IPW baseline population 1, Figure S2 (N=67,346)
p-value for interaction between lockdown, gender, and pre-existing depressive symptoms (wave 1-8): QoL (p<0·001), mental well-being (p<0·001), and loneliness (p<0·001)

**Figure S4.** Sensitivity analysis including participants who completed DNBC-18 in year 2019 and period 2 or 3. Regression of changes in QoL, mental well-being, and loneliness from pre- to during lockdown (longitudinal setup)

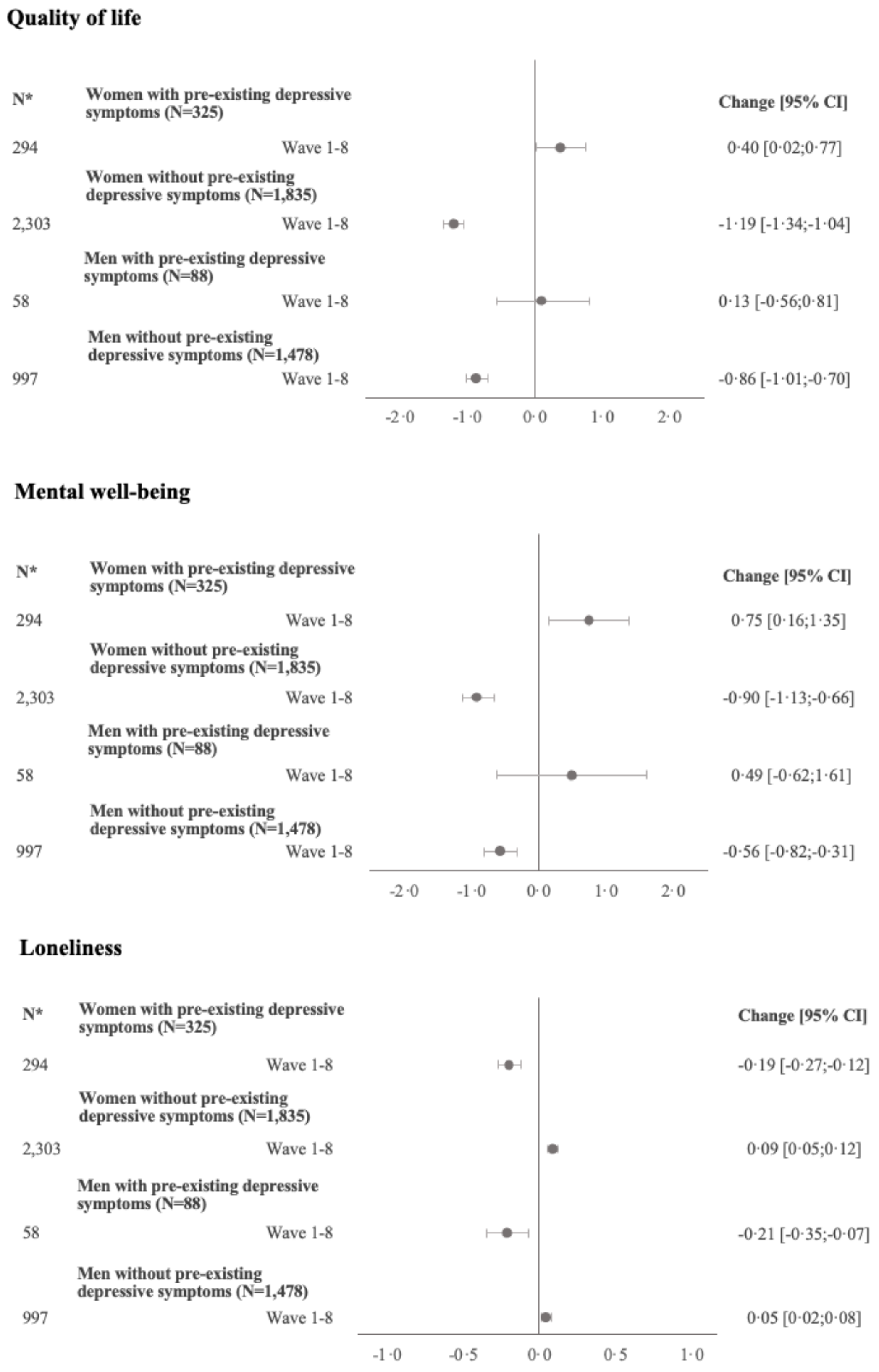


*Repeated measures
Random effect estimates and 95% CI presented (N=3,726)
(Total number, including repeated measures N=7,378)
All models were weighted by IPW baseline population 1, Figure S2 (N=67,346)
p-value for interaction between lockdown, gender, and pre-existing depressive symptoms (wave 1-8): QoL ($p<0{\cdot}001$), mental well-being ($p<0{\cdot}001$), and loneliness ($p<0{\cdot}001$)
